# A DNA intercalating dye-based RT-qPCR alternative to diagnose SARS-CoV-2

**DOI:** 10.1101/2020.12.16.20246678

**Authors:** Federico Fuchs Wightman, Micaela A. Godoy Herz, Juan C. Muñoz, José N. Stigliano, Laureano Bragado, Nicolas Nieto Moreno, Marcos Palavecino, Lucas Servi, Gonzalo Cabrerizo, José Clemente, Martín Avaro, Andrea Pontoriero, Estefanía Benedetti, Elsa Baumeister, Fabian Rudolf, Federico Remes Lenicov, Cybele C. Garcia, Valeria Buggiano, Alberto R. Kornblihtt, Anabella Srebrow, Manuel de la Mata, Manuel J. Muñoz, Ignacio E. Schor, Ezequiel Petrillo

## Abstract

Early detection of the severe acute respiratory syndrome coronavirus 2 (SARS-CoV-2) has been proven crucial during the efforts to mitigate the effects of the COVID-19 pandemic. Several diagnostic methods have emerged in the past few months, each with different shortcomings and limitations. The current gold standard, RT-qPCR using fluorescent probes, relies on demanding equipment requirements plus the high costs of the probes and specific reaction mixes. To broaden the possibilities of reagents and thermocyclers that could be allocated towards this task, we have optimized an alternative strategy for RT-qPCR diagnosis. This is based on a widely used DNA-intercalating dye and can be implemented with several different qPCR reagents and instruments. Remarkably, the proposed qPCR method performs similarly to the broadly used TaqMan-based detection, in terms of specificity and sensitivity, thus representing a reliable tool. We think that, through enabling the use of vast range of thermocycler models and laboratory facilities for SARS-CoV-2 diagnosis, the alternative proposed here can increase dramatically the testing capability, especially in countries with limited access to costly technology and reagents.

## INTRODUCTION

Since its emergence in Wuhan during December 2019, the severe acute respiratory syndrome coronavirus 2 (SARS-CoV-2) has spread worldwide, affecting health, jobs, education and social interactions of millions as well as countries’ economies ^1^. Government authorities across the world have chosen different approaches to deal with this situation, often based on social distancing and early detection followed by contact tracing. For the latter, and considering the World Health Organization’s recommendations ^1,2^, the testing capacity has turned out to be a limiting factor in numerous countries.

Given the prevalence of transmission by individuals which are asymptomatic, pre-symptomatic or suffering from mild symptoms ^3–6^, expanding the testing capacity becomes critical in order to rapidly detect and contain outbreaks. Initially, a testing strategy was adopted worldwide, which combined column-based RNA extraction from naso-pharyngeal swabs with retro-transcription followed by quantitative PCR (RT-qPCR) using TaqMan fluorescent probes. However, the whole pipeline is expensive, labor-intensive and requires specific qPCR equipment in terms of laboratory facilities. These issues, combined with the dramatically increased demand in the early days of the pandemic, resulted in testing bottlenecks that substantially delayed the tracking and isolation of infected individuals. In peripheral countries, these shortcomings tend to be amplified by economic struggling, shortage of key reagents in the international market and limited laboratory and equipment capacities. With that in sight, diagnosis alternatives have started to emerge, such as loop-mediated isothermal amplification (LAMP)-based assays, which offers a rapid diagnosis when compared to TaqMan-based RT-qPCR and does not require specialized detection equipment ^8,9^. Each alternative has different advantages and limitations, such as laboratory infrastructure requirements for receiving and fractioning viral-containing swabs, analytical sensitivity and convenience or necessity for multiplex-capable thermocyclers.

In this study, we describe the optimization of a series of custom and commercial DNA-intercalating dye alternatives based on a previously described amplicon ^10^, including specific reaction conditions for SARS-CoV-2 detection by RT-qPCR. Furthermore, we show here that this methodology is as sensitive and reliable for diagnosis as TaqMan-based RT-qPCR standards and can be used in any qPCR thermal cycler.

## Materials and methods

### Sample collection and fractionating

Swab samples were received by the Instituto de Investigaciones Biomédicas en Retrovirus y SIDA (INBIRS, CABA, Argentina) and fractionated in a Level II Biosafety Laboratory. Anonymized and randomized samples were either inactivated and transported to be extracted at Instituto de Fisiología, Biología Molecular y Neurociencias (IFIBYNE, CABA, Argentina) or directly extracted on site (as described).

### Total RNA extraction

RNA was extracted out of 300 µl of swab samples using a Chemagic 360-D automated extraction equipment (Perkin-Elmer) according to the manufacturer’s instructions.

### Retro-transcription from total RNA

Unless specified otherwise, retro-transcription reactions were done using MMLV-RT following manufacturer instructions (Invitrogen). A time-saving protocol was developed (thoroughly described in Supplementary Table 1). Program: 37°C-20’; 70°C-15’, ran in a BioRad T100 thermal cycler. For the qPCR, obtained cDNAs were diluted 1:10.

To ensure that reverse transcriptases (RT) from other providers (MMLV-RT and GoScript, Promega) could be alternatively used, we tested them in parallel using the same retro-transcription program described above. In addition to these international alternatives, we also evaluated a locally produced commercial RT, FLYE-Ultra Highway (INBIO Highway, Cat. N° K1610). It is important to note that the program’s temperatures had to be adapted for this last alternative: 60°C-20’; 95°C-15’.

### Primer design and recollection

Since unspecific priming and/or primer dimer formation are the biggest challenge to overcome when switching from TaqMan to unspecific DNA dye as the method of detection in qPCR, we screened a vast set of custom designed and published primers ^10^. Custom designed oligos were obtained using Beacon Designer Software (Premier Biosoft International, Palo Alto, CA, USA) or Primer3Plus ^11^. All candidate pairs were checked for dimer formation in both Thermo Fisher Primer Analyzer and Beacon Designer. Off-targets were analyzed using Primer Blast ^12^. Finally, the presence of common described mutations on the target sequence and cross-priming with other coronaviruses was assessed with custom R scripts. We selected 10 candidates, depicted in Supplementary Table 2, for further screening.

On the other hand, we selected two pair of primers targeting housekeeping human genes (POLR2A and U1) commonly employed in our laboratory to be used as internal controls. Their primer sequences can also be found in Supplementary Table 2.

### Primer screening

We screened the different primer pairs using a custom formulation for the PCR mix, containing SYBR^®^ Green (Invitrogen™ SYBR™ Green I Nucleic Acid Gel Stain, 10,000X concentrate in DMSO, S7563 or SYBR Green I nucleic acid gel stain 10.000X, S9430, Sigma) as the intercalating dye. Given that we expected these initial conditions to be a priori sub-optimal in terms of specificity and efficiency, several minor adjustments were made (program settings, Magnesium/DMSO concentration, among others) to obtain final working conditions (Supplementary Table 1). For the screening, we used every primer pair and performed standard curves with serial dilutions (1:4) from pooled positive samples (previously diagnosed using a commercial RT-qPCR TaqMan Kit) and evaluated absolute cycle threshold (Ct) values, reaction efficiency, correlation between expected and observed Cts, the number and position of peaks in the melting curves and potential maximum threshold for detection due to unspecific amplification.

For the internal positive control of human RNA detection, serial standard curves were performed (5 points, 1:2 from a pool of samples) to corroborate the selected housekeeping genes worked efficiently and free of non-template amplification. The custom-made mix and the INBIO Highway master mix were used for this purpose (see ahead for specifics on those mixes).

### TaqMan OneStep RT-qPCR

Following RNA extraction, detection of SARS-CoV-2 genetic material was performed using a commercial TaqMan RT-qPCR Kit (DisCoVery Kit by AP Biotech, Ref: APB-COV19R2ROX). The procedure was done following the manufacturer’s indications (https://apbiotech.com.ar/pub/media/Content/Datasheet/APB-COV19R2ROX.pdf). Alternatively, the GeneFinder RT-qPCR Kit was used for some experiments (REF: IFMR-45, https://www.fda.gov/media/137116/download). All reactions were run in a BioRad CFX-96 thermocycler.

For the comparison between the published Charité protocol ^7^ and our modified method with SYBR Green, we prepared a mix (Supplementary Table 1) based on Thermo Fisher/Invitrogen SuperScript III OneStep RT-PCR System with Platinum® Taq DNA Polymerase. The qPCR was performed according to manufacturer’s instructions: 55°C-10’; 94°C-3’; (94°C-15’’ & 58°C-30’’) 45x. All reactions were run in either an ABI Quantstudio 3 or an ABI StepOnePlus.

### SYBR Green OneStep RT-qPCR

The adapted Charité protocol for SYBR Green mix (Supplementary Table 1) was based on Thermo Fisher/Invitrogen SuperScript III OneStep RT-PCR System with Platinum® Taq DNA Polymerase. The qPCR program used: 60°C-10’; 95°C-3’; (95°C-15’’ & 60°C-40’’) 40x, melting curve. All reactions were run in either an ABI Quantstudio 3 or an ABI StepOnePlus.

### SYBR Green qPCR

All SYBR Green based mixes are thoroughly described in Supplementary Table 1. For every reaction the qPCR cycling was done with a 95°C-15’’ denaturation step and 40 cycles of 60°C-40’’ annealing/elongation, followed by a melting curve. The activation time was adapted to the one recommended by the manufacturers of each mix or Taq Polymerase (Supplementary Table 1). The master mixes tested were: INBIO Highway qPCR Mix (Ref M130, https://www.inbiohw.com.ar/es/productos/178591/pcr/Master-Mix-qPCR-con-SYBR.html), Solis Biodyne 5x HOT FIREPol EvaGreen® qPCR Mix Plus and Biodynamics Real Mix Ref B124-100. Two custom-made mixes were prepared using different Taq Polymerases (INBIO Highway T-FREE Taq REF E1202, https://www.inbiohw.com.ar/es/productos/168157/pcr/Kit-T-Free-ADN-polimerasa.html, and Qiagen Hot Start Taq REF 203203).

### Determination of the Limit of Detection (LOD)

A SARS-CoV-2 RNA standard was kindly provided by INEI-ANLIS Dr. Carlos G. Malbrán (Buenos Aires, Argentina). This standard was quantified (4×10^6^ molecules per µL) using the SARS-like Wuhan, Iv-RNA E gene Standard; 1×10^8^ copies/µL provided by the Pan American Health Organization / World Health Organization. Serial dilutions (1:10) were done, starting from a quantity of 4×10^5^ molecules per µL and ending in 4 molecules per µL. Five µL of each dilution were used in each reaction (as described before).

### Generation of a positive control for RT-qPCR

The positive control RNA was obtained by cloning a region of SARS-CoV-2 genome in the plasmid pBluescript under the control of a T7 RNA polymerase promoter. To this end, pBluescript KS^+^ plasmid was digested with BamHI and XhoI and gel-purified with an extraction kit. In turn, the insert was generated by RT-PCR using samples of positive patients as a source of SARS-CoV-2 genomic material. The primers (listed in Supplementary Table 2) included restriction enzyme adapters (BamHI and XhoI within the forward and reverse primers respectively) to allow cloning within the pre-digested pBluescript. Alternatively, the insert can be generated by gene synthesis or PCR amplification of overlapping primers. The amplified region must include the target sequence of the primers listed as RdRP. The PCR product was subsequently digested with previously mentioned enzymes and ligated to the linearized vector with T4 DNA ligase. The cloned products were transformed into DH5α E. coli, followed by standard plasmid miniprep protocols. The plasmid constructs obtained were linearized with XhoI restriction enzyme and incubated with T7 RNA polymerase for in vitro transcription. Finally, DNA in the reaction was degraded by incubation with RNAse-free DNase I according to the manufacturer’s indications. Total RNA was purified with AGENCOURT® RNAClean® XP magnetic beads (Beckman Coulter) and quantified with a Nanodrop (Thermo Scientific) to be used as a positive control RNA standard sample in RT-qPCR reactions.

## Results

### Selection of a primer pair for SARS-CoV-2 detection

An initial screening was conducted with a set of candidate primer pairs (Supplementary Table 2). We tested the different primer pairs on serial dilutions of a pool of positive samples (already diagnosed using a commercially available TaqMan Kit) and no-template controls. We scored the Ct value for each of these reactions and assessed the different primers for: amplification efficiency, R^2^ for the linear regression of Ct vs. log(quantity), melting curves and the amplification cycle at which unspecific PCR products start to be detected in the no-template controls. Figure 1A shows the amplification and melting curves for each primer pair, and Figure 1B a summary of the data obtained. Based on this information, the RdRP primer pair was chosen for further studies. Furthermore, an *in vitro* transcribed RNA was generated and used as a positive control to monitor the quality of each RT-qPCR run. This is particularly relevant when using custom-made mixes that, according to the preparation, could yield variable outcomes.

**Figure 1.**
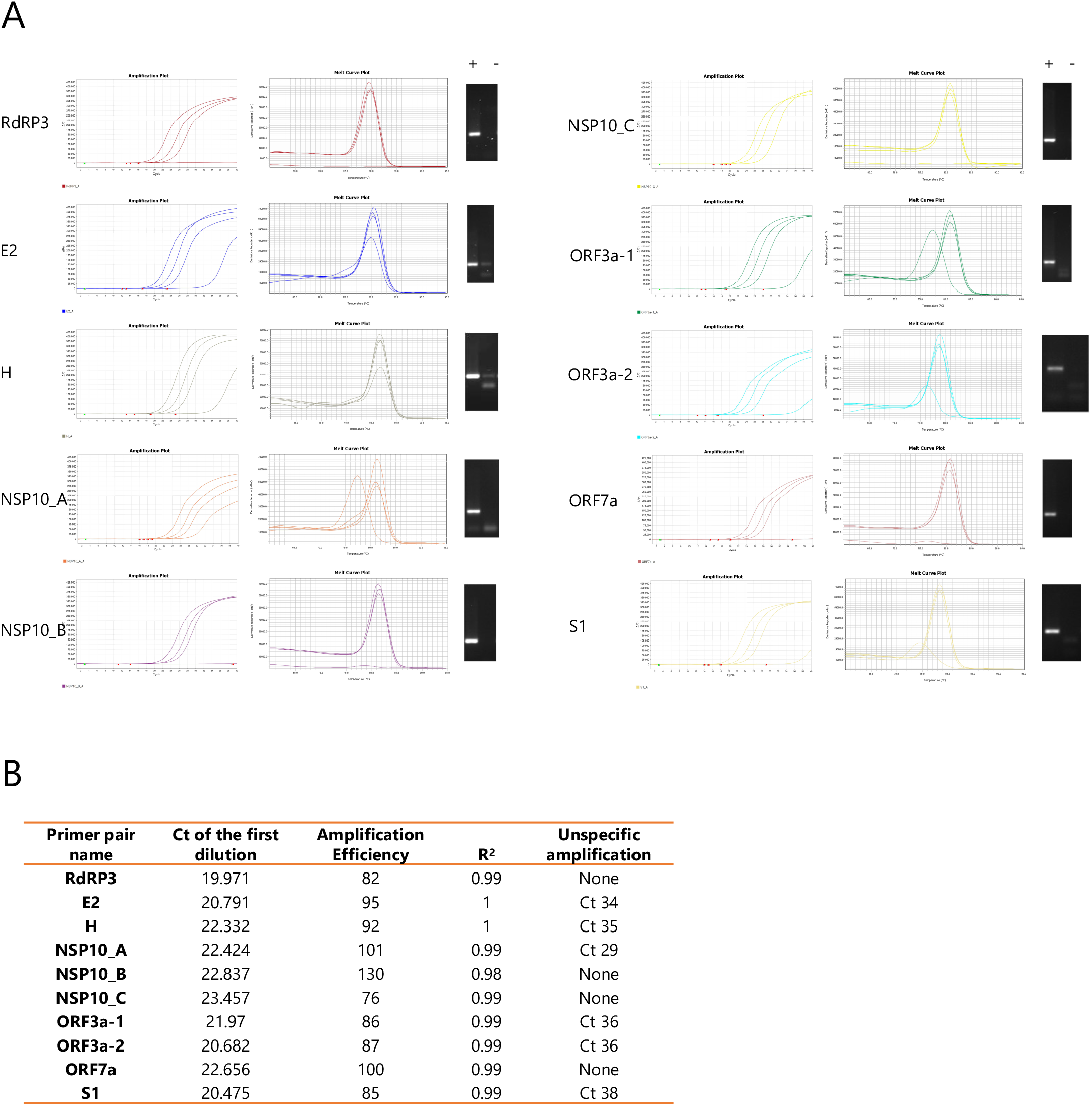
Selection of a primer pair for SARS-CoV-2 detection. (A) Serial dilutions of a pool of positive samples for each primer pair. Representative amplification (*left panel*) and melting curves (*middle panel*) are shown. Corresponding RT-qPCR products were solved by gel electrophoresis and representative images of positive samples (+) and negative controls (-) are shown (*right panel*). (B) Summary of the data generated for selection of a highly specific primer pair. *Ct*: cycle threshold. *Amplification efficiency* and *R*^*2*^ relates to the log-linear fit of the relation between *Relative quantity vs. Ct* for known serial dilutions of a positive control. An amplification efficiency of 100% represents a decrease of one Ct for a 1:2 dilution. *Unspecific amplification*: *Ct* of amplification curves observed in negative controls (without template), probably due to primer dimers.

### SYBR green RT-qPCR as a suitable alternative to TaqMan RT-qPCR for SARS-CoV-2 detection

With the goal of providing experimental alternatives to the gold standard TaqMan RT-qPCR, several qPCR mix options were tested. These all relied on either SYBR Green or Eva Green, as alternative intercalating dyes, for the detection of the genetic material and ranged from one-step reaction mixes to two-step methods using a custom mix (see Materials and Methods). The main objective was to facilitate a range of customizable solutions for specific needs that might arise during the COVID-19 diagnosis pipeline. Additionally, we aimed at sorting them from the most timesaving to the least, irrespectively of their cost and/or availability in the local market. To cope with the main disadvantage of unspecific amplification observed when using intercalating dyes instead of labeled probes, we defined the critical amplification cycle at which unspecific PCR products were detected and set it as the cutoff cycle of our methods’ detection limit. While we are conscious that this strategy impairs the limit of detection (LOD) marginally, it seemed more reasonable to prioritize specificity in this context in order to diminish the occurrence of false positives.

Firstly, we adapted the Charité protocol published by the World Health Organization (WHO) to use SYBR Green instead of TaqMan probes. For this purpose, we evaluated the pair of RdRP primers selected from the screening, added the dye and adjusted the cycling program and mix composition (see Methods). Interestingly, there was a substantial increase in sensitivity when using the intercalating dye compared to the TaqMan probes N and ORF1ab (Figure 2A), with an average decrease in Cts of approximately 4 cycles, which allowed for every true positive sample to be detected as positive. When comparing the limit of detection (LOD) of a SARS-CoV-2 standard RNA (see Methods), we found higher sensitivity when using SYBR Green rather than a TaqMan-based approach with an amplicon in the same region of the viral RNA (using RdRP primers and probe P1 ^7^) (Figure 2B). We selected a Ct cutoff of 35, since it ensured full specificity (Figure 2C and Supplementary Data 1). Consequently, the LOD for the SYBR Green-based RT-qPCR with the RdRP primer pair is approximately 20 copies of viral genome (Figure 2B). Since 5 µl of RNA are used per reaction, this means that the intercalating dye method could detect as low as 4 copies (c) of viral RNA per µl or 4000 c/ml.

**Figure 2.**
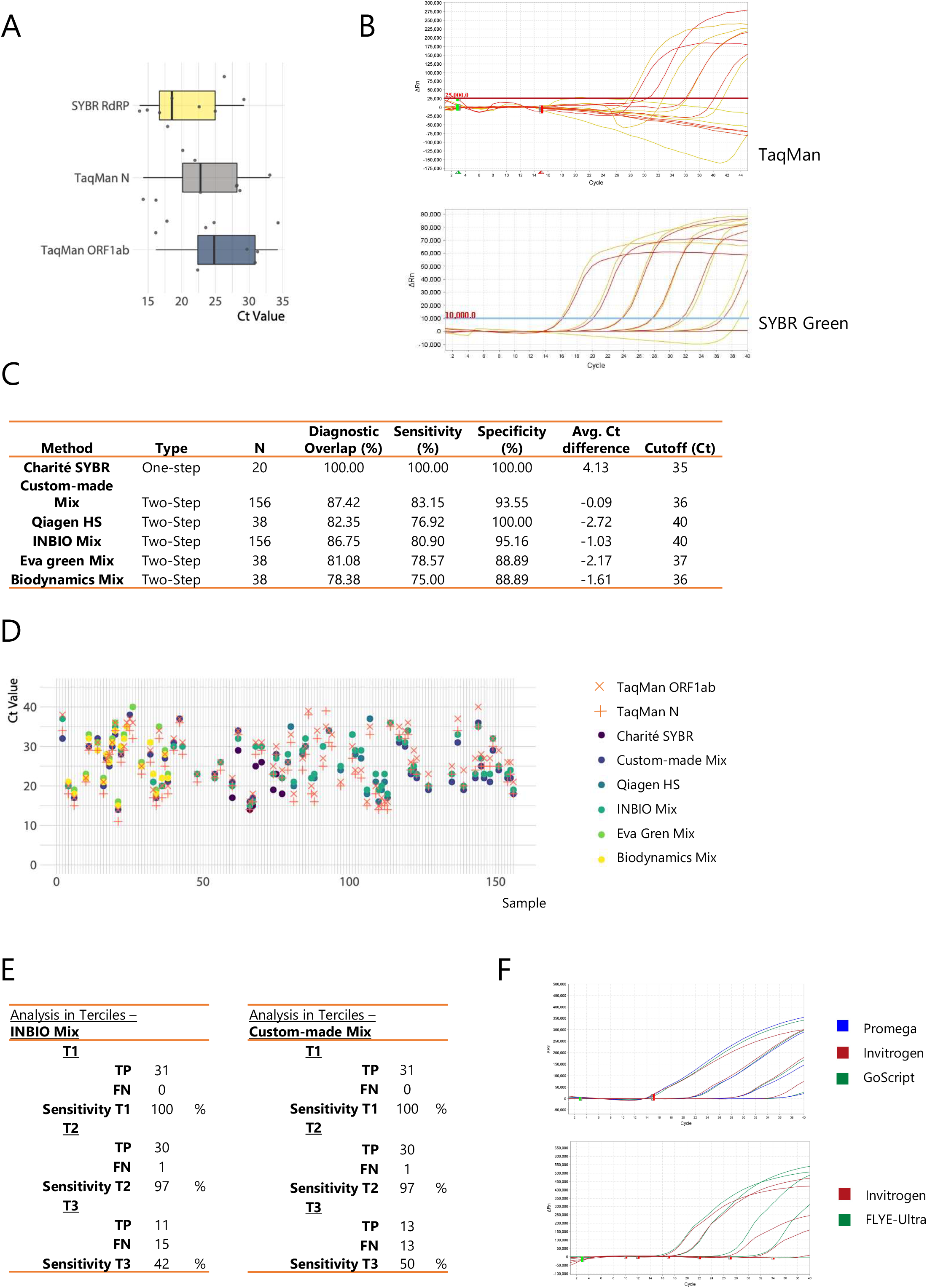
Using an intercalating dye can yield similar sensitivity and specificity as TaqMan probes. (A) Boxplot of the Ct shift for the SYBR Green RT-qPCR (RdRP amplicon) versus two different TaqMan-based reactions (N and ORF1ab amplicons) using the *Charité* protocol. (B) RT-qPCR amplification curves for serial dilutions of a SARS-CoV-2 standard RNA, from 4×10^5^ copies per µL (c/µL) to 4 c/µL, using either the unmodified *Charité* protocol (TaqMan) or the one adapted for an intercalating dye (SYBR Green). (C) Summary of the performance obtained using the different commercial and customized RT-qPCR protocols evaluated. (D) Ct values for each sample (arbitrarily numbered) tested with every RT-qPCR protocol developed and optimized in this work. Not all samples were evaluated with all protocols. Missing points represent negative (not amplified) samples. (E) A drop in diagnostic sensitivity using two-step RT-qPCR approaches is circumscribed to samples with higher Ct values. Table summarizing the sensitivity observed using either the INBIO master mix or the custom-made mix when separating the positive samples in terciles (T1-T3) according to the previously assessed Ct value using the RT-qPCR DisCoVery kit (ORF1ab and N amplicons). (F) RT-qPCR amplification curves for either serial dilutions of a pool of positive samples (*top panel*) or random samples (*bottom panel*), using four different retro-transcriptases for cDNA synthesis.

Next, we decided to switch to a classic two-step RT-qPCR protocol to broaden the possibilities of reagents to be used for the procedure. We tested the different qPCR mixes in a panel of samples (38 - 156 samples) comparing in parallel with a TaqMan diagnosis commercial kit (DisCoVery by AP-Biotech) as our gold-standard. By analyzing the overlap with this gold-standard and calculating the corresponding sensitivity and specificity values, we show that every commercial qPCR master mix we tried (INBIO Highway Ref M130, Solis Biodyne 5x HOT FIREPol EvaGreen® qPCR Mix Plus and Biodynamics Real Mix Ref B124-100 see Methods for details) and two custom mixes (INBIO Highway Taq E1202 preparation and Qiagen Hot Start Taq 203203 see Methods for details) proved efficient for diagnosis purposes (Figure 2C and Supplementary Data 1) with the proposed primers and qPCR program, an adjusting a specific Ct cutoff for each case.

Remarkably, none of the mixes showed an average Ct shift higher than 3 cycles (Figures 2C-D and Supplementary Data 1) when comparing with the DisCoVery SARS-CoV-2 kit. As expected, the decrease in sensitivity in the 2-step options is biased towards the samples with low amounts of viral RNA, as measured by the DisCoVery SARS-CoV-2 RT-qPCR kit (Figure 2E). While this would result in an increase of the number of false negatives, it would affect people with lower viral load specifically, which tend to correlate with lower transmission ^13,14^. Furthermore, we propose that this is a minor drawback if it is counterbalanced by a large increase in the testing capabilities due to the higher availability and lower costs of reagents and the use of more qPCR thermocyclers.

Lastly, four different reverse transcriptases were tested, and the RT procedure was simplified and streamlined to be time saving and to reduce manipulation of samples (see Methods). Our results showed that all the tested RT enzymes worked equally well (Figures 2F and G).

### Selection of a human housekeeping gene to be used as an internal control

Two primer pairs, targeted to human POLR2A mRNA and U1 snRNA, were shown to work adequately with the proposed mixes (Figures 3A-B). Thus, either one can be used as an internal control for diagnosis.

**Figure 3.**
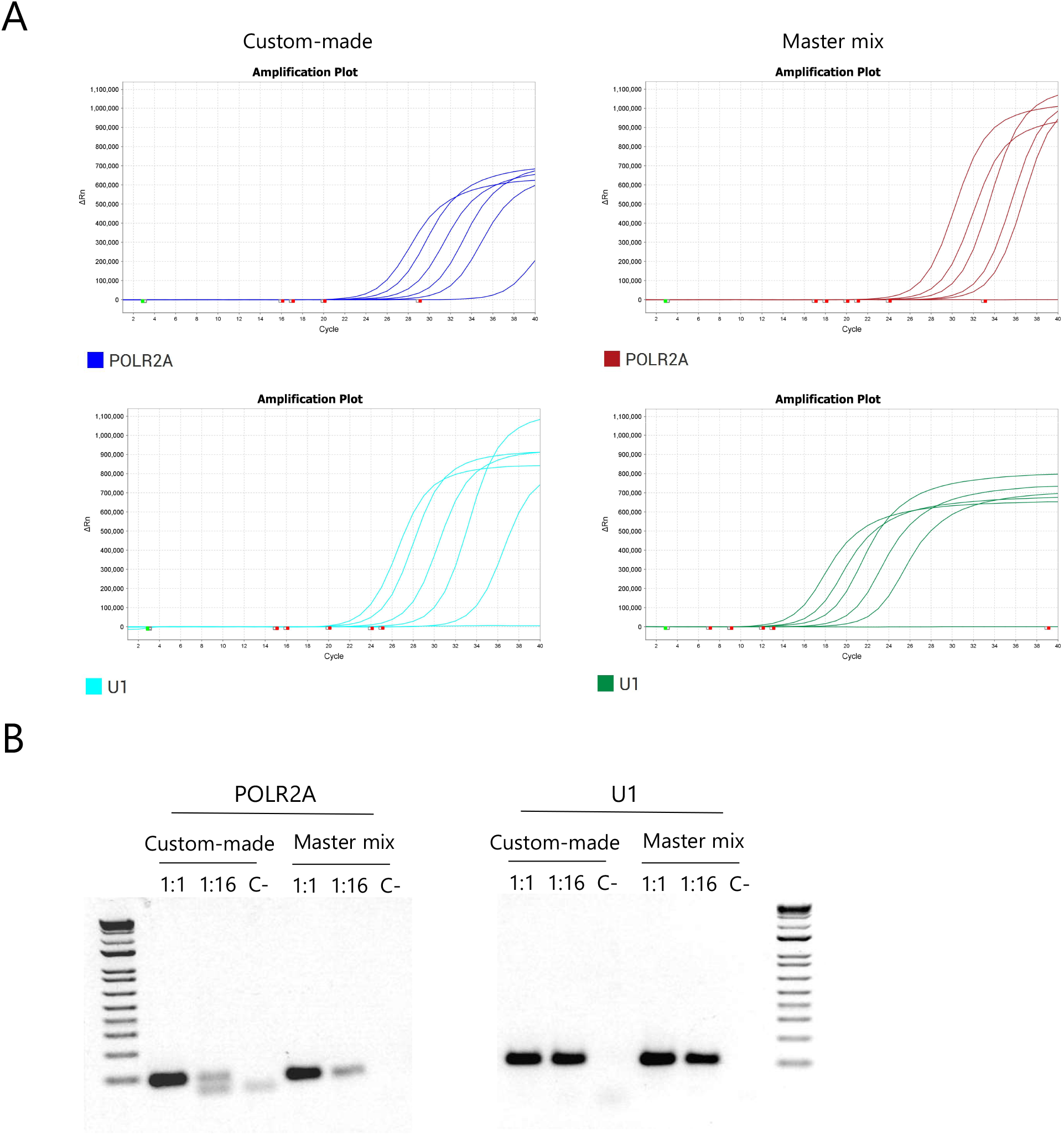
Selection of a human housekeeping gene to be used as internal control. (A) Representative amplification curves for POLR2A and U1 using both the custom-made mix and the INBIO master mix. (B) Agarose gel electrophoresis of the products obtained after qPCR of 1:1, 1:16 dilutions and the negative control respectively, for each primer pair using both mixes.

## Discussion

The aim of this report is to provide alternative RT-qPCR approaches for diagnosis of COVID-19 caused by the severe acute respiratory syndrome coronavirus 2 (SARS-CoV-2). We present alternative strategies to the current gold standard (TaqMan-based RT-qPCR), mainly based in intercalating dye-based detection. The key advantages of intercalating dyes (particularly SYBR Green) are their wide availability, low cost, simplicity of use, wide range of thermocyclers capable of detecting them and the and the easy ordering of oligonucleotides without the requirement for labeled probes.

In that line, probably the most time-saving option that we favor would be to follow the Charité protocol adapted for SYBR Green. Nevertheless, we decided to optimize different alternatives in case that One-Step master mixes would appear out of reach either for their cost or due to unavailability in local or regional markets. This might sound far-fetched in economically strong countries but needs to be deeply considered for those peripheral countries struggling with recession, shortages and lack of infrastructure. Consequently, we generated Two-Step protocols that can be followed with commonly used reverse transcriptases (MMLV-RT) from different vendors and a variety of qPCR mixes. For the former we tried four different RT brands, all of which performed equally well, leading to the speculation that RT from additional vendors could probably be used as well, after little experimental validation. Similarly, as long as the proposed qPCR cycling program and pair of primers are used, all the qPCR mixes we tested, including a very accessible custom formulation, proved to be useful for the described procedure.

The main limitation we encountered when switching to intercalating dyes for detection was unspecific amplification. Unlike labeled probes, which have an extra layer of specificity, negative samples analyzed with SYBR Green-based qPCRs may sometimes exhibit unspecific sigmoidal fluorescence curves with high Ct values due to, for example, primer dimer amplification. While the wide screening of primer pairs that we performed resulted in an amplicon where this issue was greatly diminished, it came at the cost of analyzing and selecting a specific threshold to consider a sample positive for each qPCR mix. The thresholds were selected to exclude Cts arising from occasional amplification of negative controls (lacking any template). As described above, this did not have a major effect on the sensitivity of detection. It is worth mentioning that a negative result on this kind of test does not rule out completely the possibility of actual SARS-CoV-2 infection. Other variables, like the progression of the disease at the moment of the swab or the quality of the sample, also influence the final result of the test. A proper diagnosis must take into account all these variables and not only the result of the qPCR.

The work presented here clearly broadens the possibilities regarding the use of reagents and equipment. Furthermore, this method based on intercalating dyes displays similar diagnostic power to widely used qPCRs based on labeled probes. Hence, combining these alternatives for SARS-CoV-2 detection by RT-qPCR with simple and accessible clinical sample and RNA preparation methods, would render a significant improvement on testing capacities, specially of low-to-medium income countries.

## Data Availability

All data referred to in the manuscript is part of the main figures or the suppl. information. If there is any other data a reader may need or further info on methods, do not hesitate to contact the authors.

## Acknowledgments

We thank Alejandro Colman Lerner, Andrea Barta, Alwin Köhler (Vienna COVID-19 Detection Initiative) and Rodrigo A. Gutiérrez for helpful feedback and help. We are deeply grateful for the constant support provided by the whole community at IFIBYNE, INBIRS and INEI-ANLIS-Malbrán even under unprecedented circumstances. Without their dedication and commitment, this research would have not been possible. We specially acknowledge the help of Rocío S. Tognacca, Luciano Marasco and Nicolás Gaioli.

## Funding

This work was supported by the Agencia Nacional de Promoción Científica y Tecnológica (ANPCyT, grant IP-COVID-19 429 to ARK) and by Fondos del Presupuesto del Servicio de Virosis Respiratorias, INEI-ANLIS Malbrán (MA, AP, EB and EB). FFW is an ANPCYT fellow; MAGH, JCM, JNS, LB, NNM, MP, LS and GC are fellows from Consejo Nacional de Investigaciones Científicas y Técnicas de Argentina (CONICET); JC and VB are support staff for research and development (CPA) from CONICET; FRL, CCG, ARK, AS, MdlM, MJM, IES and EP are career investigators from CONICET.

## Conflict of interest

The authors declare that they have no conflict of interest.

## Notes

### Competing Interest Statement

The authors have declared no competing interest.

### Funding Statement

This work was supported by the Agencia Nacional de Promocion Cientifica y Tecnologica (ANPCyT, grant IP-COVID-19 429 to ARK) and by Fondos del Presupuesto del Servicio de Virosis Respiratorias, INEI-ANLIS Malbran (MA, AP, EB and EB). FFW is an ANPCYT fellow; MAGH, JCM, JNS, LB, NNM, MP, LS and GC are fellows from Consejo Nacional de Investigaciones Cientificas y Tecnicas de Argentina (CONICET); JC and VB are support staff for research and development (CPA) from CONICET; FRL, CCG, ARK, AS, MdlM, MJM, IES and EP are career investigators from CONICET.

### Author Declarations

This study used the remaining volume of anonymized samples that had been collected for clinical diagnosis. Under these circumstances, and for studies involving development of COVID-19 diagnostic tools, our IRB (the Research Ethics Committee from Fundacion Huesped in Buenos Aires, Argentina) deemed unnecessary to obtain informed consent from the patients and waived the ethical approval.

